# Bicuspid Aortic Valve Disease With Early Onset Complications: Characteristics And Aortic Outcomes

**DOI:** 10.1101/2024.03.11.24304079

**Authors:** Maximilian A. Selbst, Megan K. Laughlin, Colin R. Ward, Hector Michelena, Anna Sabate-Rotes, Lisa Bianco, Julie De Backer, Laura Muiño Mosquera, Anji T. Yetman, Malenka M Bissell, Maria Grazia Andreassi, Ilenia Foffa, Dawn S. Hui, Anthony Caffarelli, Yuli Y. Kim, Dongchuan Guo, Rodolfo Citro, Margot De Marco, Justin T. Tretter, Shaine A. Morris, Kim L. McBride, Simon C. Body, EBAV Investigators, Siddharth K. Prakash

## Abstract

Bicuspid aortic valve (BAV) is the most common congenital heart malformation in adults but can also cause childhood-onset complications. In multicenter study, we found that adults who experience significant complications of BAV disease before age 30 are distinguished from the majority of BAV cases that manifest after age 50 by a relatively severe clinical course, with higher rates of surgical interventions, more frequent second interventions, and a greater burden of congenital heart malformations. These observations highlight the need for prompt recognition, regular lifelong surveillance, and targeted interventions to address the significant health burdens of patients with early onset BAV complications.

## 1. Introduction

Bicuspid aortic valve is the most common congenital heart defect, affecting 0.5-2% of the general population. Bicuspid aortic valve can be associated with significant cardiac complications including aortic regurgitation or stenosis, and thoracic aortic aneurysm predisposing to thoracic aortic dissection. The clinical spectrum of bicuspid aortic valve disease ranges from lesions that are diagnosed *in utero* or at birth to incidental findings in asymptomatic adults. In some cases, bicuspid aortic valve may be a feature of genetic syndromes such as Turner syndrome or Loeys-Dietz syndrome. Prompt diagnosis of bicuspid aortic valve and identification of patients who are at elevated risk for early onset complications is essential because treatment frequently requires intervention, frequent surveillance, and intensive medical therapies. We hypothesize that people with early onset complications of bicuspid aortic valve may have anatomic, clinical, or genetic predictors that distinguish them from the majority of individuals with bicuspid aortic valve who progress along a more benign clinical course. Therefore, we characterized a unique cohort of young adults who presented between ages 17 and 30 with moderate or severe valvular or aortic disease.

## 2. Materials and Methods

Early onset bicuspid aortic valve probands were recruited from 16 collaborating investigators who referred eligible individuals to the data coordinating center at UTHealth Houston. The study protocol was approved by the Committee for the Protection of Human Subjects at the University of Texas Health Science Center at Houston (HSC-MS-11-0185). All participants signed a written consent form, and study procedures were conducted in compliance with the ethical standards of the relevant national guidelines on human experimentation (HHS regulations 45 CFR part 46) and with the Helsinki Declaration of 1975, as revised in 2008.The study protocol was previously published.^1^ Inclusion criteria were maximum aortic Z-score (root or ascending) > 4, thoracic aortic dissection, moderate or severe aortic stenosis or regurgitation, or valvular or aortic intervention prior to age 30. Individuals with genetic syndromes or clinically discovered mutations of causal bicuspid aortic valve or HTAD genes were excluded.^1,2^ Unpaired t-tests and chi-squared tests were used to compare subgroups.

## 3. Results

A total of 279 probands were included in this analysis. The mean age at diagnosis was 18 years. Two-thirds were male and almost 30% had other congenital heart malformations, most frequently aortic coarctation, ventricular septal defects, atrial septal defects, patent ductus arteriosus, or mitral valve abnormalities. Half of participants had their first surgery in childhood (mean age 7), and 25% had their first surgery as adults (mean age 33). One-third of adult surgeries were second operations after a childhood surgery. Aortic valve repair or replacement (44%) was the single most common indication for intervention. One-quarter underwent isolated repair of the ascending aorta and 21% had combined valve and aortic operations (Bentall or David). Fifteen probands (22%) required at least one additional intervention after their index operation. Eight of the initial interventions occurred in childhood and required reinterventions before age 25. The most frequent indications for reintervention were aortic aneurysm repair and aortic valve replacement. The median time between the first and second interventions was 14 years.

We compared early onset bicuspid aortic valve demographics to a previously published meta-analysis of 4379 adult cases and a retrospective cohort study of Olmsted County residents with bicuspid aortic valve.^2,3,4^ The authors of the Olmstead County study defined a complex valvulo-aortopathy phenotype similar to the early onset bicuspid aortic valve phenotype, except for the inclusion of syndromic and complex congenital cases excluded from the early onset cohort. The results highlight the relative youth of the early onset bicuspid aortic valve cohort compared to published adult bicuspid aortic valve cohorts. The frequency of aortic regurgitation was significantly higher in early onset bicuspid aortic valve probands. The similarities between the early onset and complex-valvulo-aortopathy cohorts are also apparent (Table A).

**Table A.** Comparison of EBAV cohort to published bicuspid aortic valve cohorts. Meta: meta-analysis of 4379 adult BAV cases^3^; EBAV: bicuspid aortic valve with early onset complications requiring intervention prior to age 30; Complex: bicuspid aortic valve with complex valvulo-aortopathy phenotype, as defined by Yang *et al*. ^4^;AR: Aortic Regurgitation, mild or greater in severity; AS: Aortic Stenosis, mild or greater in severity; NA: not available.

|  | **Meta** | **EBAV** | **Complex** |
| --- | --- | --- | --- |
| **Age at presentation (y)** | 52±7 | 18±12 | 14±12 |
| **Male (%)** | 67±14 | 67 | 63 |
| **AS (%)** | 49±24 | 31 | 25 |
| **AR (%)** | 28±18 | 38 | 47 |
| **AS+AR (%)** | 20±17 | 11 | NA |
| Family member with BAV (%) | 6.4% | 32 | NA |

Right-non-coronary cusp fusion was more prevalent in early onset bicuspid aortic valve probands (32%) than in community-living adults with bicuspid aortic valve (14%).^5^ Previous data corroborates our observations that aortic regurgitation is more pronounced in young individuals with unusual valve morphologies.^6^ The proportion of early onset bicuspid aortic valve probands who had at least one relative with bicuspid aortic valve (32%) was significantly higher than previous estimates (5-10%).^7,8^ These observations underscore the increased burden of bicuspid aortic valve-related complications and signal the probable enrichment of highly penetrant genetically triggered disease in the early onset bicuspid aortic valve cohort.

## 4. Discussion

The overall goal of this study was to investigate the health burden of people with early complications of bicuspid aortic valve disease. The mean age at presentation of the early onset bicuspid aortic valve cohort was 18 years. In contrast, the mean age at presentation of adult bicuspid aortic valve patients from 15 prospective studies was 52 years.^3^ Only 14% of participants in the largest natural history study of adult bicuspid aortic valve disease were less than 30 years old at presentation.^5^ Thus, the early onset bicuspid aortic valve cohort is an extreme phenotype sample of bicuspid aortic valve disease. We hypothesized that early diagnosis may predict a more severe disease course due to additional congenital heart malformations and more frequent reintervention. We confirmed that early onset bicuspid aortic valve participants did require surgical interventions at a higher rate, underwent second interventions more frequently within 15 years of their index procedures, and had more frequent secondary congenital malformations than older bicuspid aortic valve patients that comprise the majority of longitudinal BAV study cohorts.^5,9^ The proportions of females (33%) and family members with a bicuspid aortic valve (32%) were substantially higher than previously reported estimates, validating our hypothesis that the penetrance of genetically triggered disease may be increased in early onset bicuspid aortic valve disease.^10,11^ Most participants who required more than one procedure had the initial aortic repair in childhood and underwent subsequent aortic valve repair or replacement before age 25. The necessity of repeated surgical interventions in such a young population has significant implications for life expectancy and quality of life. Moreover, the long-term durability of valve interventions in this population remains uncertain, raising the possibility that they may require future interventions. In the Olmstead County study, the subgroup of individuals with complex-valvulo-aortopathy, defined similarly to early onset bicuspid aortic valve disease, were found to have decreased long-term survival.^3^ Our observations highlight stark contrasts between the prognosis of the early onset bicuspid aortic valve cohort and late onset bicuspid aortic valve disease that becomes clinically apparent after age 50, highlighting the need for prompt recognition, regular surveillance, and targeted interventions to address the significant health burdens of early onset bicuspid aortic valve patients. This study also demonstrates the potential value of familial screening or genetic testing to identify relatives who may be at risk for bicuspid aortic valve-related complications.

The early onset bicuspid aortic valve cohort represents a small subset (10-15%) of bicuspid aortic valve cases, even with our multicenter approach, and we acknowledge the limitations related to subgroup analysis of a rare phenotype.

## Data Availability

All data produced in the present study are available upon reasonable request to the authors.

https://www.redcap.com

## Abbreviations

BAV: Bicuspid Aortic Valve
HTAD: heritable thoracic aortic disease

## 5. Acknowledgements

The authors are indebted to Jacqueline Jennings, Joana Castillo, and Kayla House for enrollment and data curation. The EBAV Investigators are: Shaine A. Morris, Rita Milewski, Giuseppe Limongelli, Alessandro Della Corte, Laura Perrone, Yuli Y. Kim, Hector Michelena, Maria G. Andreassi, Arturo Evangelista, Denver Sallee, Anji Yetman, Kim McBride, Eduardo Bossone, Rodolfo Citro, Dawn S. Hui, Malenka M. Bissell, Andrea Ballotti, Ilenia Foffa, Margot De Marco, Anthony Caffarelli, Rita Weise, Julie DeBacker, Laura Muiño Mosquera, Robbin Cohen, Laura Dos Subira, Justin T. Tretter, Anna Sabate-Rotes, Martina Caiazza, Lamia Ait Ali, Francesca Pluchinotta, and Simon C. Body. We are also grateful to the International Bicuspid Aortic Valve Consortium (BAVCon).

## 6. Financial Support

This work was supported in part by R01HL137028 (S.P.) and R01HL114823 (S.B.).

## Conflicts of Interest

None.

## Ethical Standards

The authors assert that all procedures contributing to this work comply with the ethical standards of the relevant national guidelines on human experimentation (HHS regulations 45 CFR part 46) and with the Helsinki Declaration of 1975, as revised in 2008, and has been approved by the Committee for the Protection of Human Subjects at the University of Texas Health Science Center at Houston.

## Notes

### Competing Interest Statement

The authors have declared no competing interest.

### Clinical Protocols

https://www.ncbi.nlm.nih.gov/pmc/articles/PMC7549564/

### Funding Statement

This work was supported in part by R01HL137028 (S.K.P.) and R01HL114823 (S.C.B.).

### Author Declarations

The study protocol was approved by the Committee for the Protection of Human Subjects at the University of Texas Health Science Center at Houston (HSC-MS-11-0185). All participants signed a written consent form, and study procedures were conducted in compliance with the ethical standards of the relevant national guidelines on human experimentation (HHS regulations 45 CFR part 46) and with the Helsinki Declaration of 1975, as revised in 2008. IRB of the University of Texas Health Science Center at Houston (HSC-MS-11-0185) gave ethical approval for this work.

